# Adverse Childhood Experiences Reorganise the Brain-Personality Network Across the Psychosis Spectrum

**DOI:** 10.64898/2026.06.15.26354444

**Authors:** Pierfrancesco Sarti, Giacomo Cecere, Hans-Latif Hua Dällenbach, Roya Hüppi, Akhil Ratan Misra, Victoria Edkins, Wolfgang Omlor, Johanna M.C. Blom, Werner Surbeck, Philipp Homan

## Abstract

Exposure to adverse childhood experiences is a pervasive risk factor for psychosis, exhibiting a linear relationship across the psychosis spectrum from subclinical schizotypal traits to schizophrenia spectrum disorders. While this association is often conceptualised within the vulnerability-stress framework, the systemic mechanisms through which childhood trauma reconfigures the brain–personality interactome remain poorly understood. We examined clinical, neuropsychological, and neuroimaging data from a sample of low-and high-schizotypy individuals, and patients with a diagnosis of schizophrenia spectrum disorder (N=120). Our aim was to map how trauma reconfigures interactions between neurobiology and schizotypal phenomenology. We adopted a mixed graphical model approach to jointly estimate conditional dependencies between childhood trauma, regional brain morphometry, and schizotypal traits across the psychosis spectrum. Our results show that childhood trauma reconfigures the brain-personality network, shifting it from a state driven by cognitive processes to one anchored in emotional (limbic) reactivity. This transition is marked by the increased influence of impulsive traits and a significant strengthening of connections within the salience network. These changes converge with a reduced thickness of the frontal executive regions, the brain’s ‘control centres’, identified in our models. Collectively, our results suggest a structural–phenomenological decoupling, where trauma-conditioned affective circuits may bypass weakened top-down regulatory controls. These findings highlight the necessity of using integrative frameworks to capture how trauma fundamentally reshapes the relationship between the brain and schizotypal personality.

## 1. INTRODUCTION

Adverse childhood experiences (ACEs) are among the most potent environmental risk factors implicated in the development of psychotic phenomena^1–3^. Meta-analytic evidence reveals a dose-response relationship between exposure to childhood abuse or neglect and the incidence of schizophrenia spectrum disorders (SSDs)^3,4^. This association extends to schizotypy, a multidimensional constellation of subclinical personality traits that mirrors the phenomenology of clinical psychosis^5^. In this population, childhood trauma has been associated with intensified symptom expression, reduced functional capacity, and poorer clinical outcomes^5^. Traditionally, these links are often conceptualised within the framework of the vulnerability-stress model, according to which the development of psychosis results from an interaction between an individual’s underlying biological diathesis and the progressive accumulation of environmental stressors^6,7^. ACEs occur during sensitive neurodevelopmental windows when large-scale neural networks supporting emotion regulation, social cognition, and executive control are undergoing maturation ^5,8^. Within the vulnerability-stress framework, early adversity may act as a biological disruptor of neuroendocrine and immune signalling, triggering widespread, coordinated changes, most notably excessive synaptic pruning during adolescence^9,10^, capable of altering the development and organisation of neural circuits underlying core cognitive and affective processes^11^. Such trauma-related reorganisation may lead to enduring changes in brain structure and function that, in turn, shape the emergence and severity of schizotypal traits and increase vulnerability to psychotic experiences across the lifespan^1–3^.

Neuroimaging research has identified structural alterations in prefrontal, insular, limbic, and cingulate regions that are associated with traumatic experiences^12–14^. These same regions feature prominently in structural and functional models of schizophrenia^15^ and as candidate neuroimaging biomarkers of psychosis risk^16–18^.

Recent findings from the ENIGMA schizotypy working group indicate that higher levels of schizotypy are associated with thicker cortex in the right middle temporal gyrus, right insula, and left caudal anterior cingulate cortex, as well as thinner left caudal middle frontal gyrus; these associations were only present in individuals exposed to higher levels of childhood trauma^19^. These findings suggest that the severity of childhood trauma experienced may impact the neurodevelopmental trajectories of brain regions implicated in affect regulation and critical for cognitive and integrative processes in relation to the expression of schizotypy^5,13,14,17,20^. However, it remains unclear how childhood adversity shapes the association between brain morphology and schizotypal and psychosis-proneness^21,22^.

Research on trauma and psychosis has historically relied on bivariate or unidirectional models that examine isolated pathways^3,23^. Over the past decade, the network paradigm has emerged as a compelling alternative, conceptualising mental health disorders as systems of mutually reinforcing elements (symptoms, cognitive processes, and environmental stressors) that causally influence each other^24–26^. This approach evaluates the dependencies between variables while also examining the system’s global architecture^27–29^. By using metrics such as overall connectivity and centrality, it provides quantitative indices of network resilience and vulnerability, identifying the system’s capacity to maintain structural integrity or functional coherence under perturbation^24,30^.

Several influential studies have applied network methods to trauma and psychosis, demonstrating that childhood trauma is linked to psychotic symptoms via specific intermediary nodes such as anxiety, depression, and dissociation, rather than through a diffuse, non-specific effect^31,32^. Subsequent analyses have mapped complex pathways between ACE domains, symptoms, and functional outcomes in first episode psychosis^33,34^. Recent work has analysed the interplay between childhood trauma and bodily self-disturbances^35,36^. These studies suggest that certain experiential anomalies may act as “bridges” linking early-life adversities to the expression of positive symptoms. At a broader conceptual level, these findings align with contemporary formulations of schizophrenia as a disorder of disconnected brain and symptom networks^2,37^.

Despite this progress, most network research remains restricted to clinical symptom dimensions. By extending this paradigm to also include individuals with schizotypal traits, it may be possible to examine how childhood trauma reshapes the architecture of associations in ways that may precede or predispose individuals to clinical illness^1,38–40^. Furthermore, structural neuroimaging measures have rarely been integrated as full nodes within network studies. Morphometric alterations are typically analysed in parallel to, rather than in interaction with, trauma and personality measures^12,17,20^. This separation obscures potential “vulnerability hubs” (neuroanatomical regions whose connectivity to schizotypal traits may be particularly sensitive to trauma exposure and thus critically positioned in the pathway from adversity to disorder).

The present study addresses these gaps by adopting a mixed graphical model approach that jointly estimates conditional dependencies between ACEs, regional brain morphometry, and multidimensional schizotypal traits across the psychosis spectrum. In other words, this approach allows us to see, in a single picture, which aspects of early trauma, brain structure, and psychosis-like experiences are directly connected to each other, and which apparent connections disappear once other factors are taken into account. Our sample includes patients who had received a diagnosis of SSD and healthy individuals with high and low-schizotypy. By treating trauma not just as a background risk factor but as something that actively reshapes the relationship between brain structure and psychosis-like experiences, we aimed to map how it reorganises those connections. To achieve this aim, we tested three hypotheses: 1) Following the vulnerability-stress model^41^, ACE prevalence and severity would increase across the psychosis spectrum gradient. 2) There would be a corresponding gradient of structural alteration, with the SSD group exhibiting the most pronounced reductions in cortical thickness and volume compared to both high- and low-schizotypy healthy individuals. 3) Most critically, childhood trauma would act as a systemic catalyst, reconfiguring the global architecture of morphometric-schizotypy associations. In other words, we anticipated that childhood trauma would fundamentally reshape the way brain structure and schizotypal traits relate to one another. This shift would manifest as the emergence of specific “vulnerability hubs” within the network, potentially altering the balance between circuits dedicated to cognitive control and those involved in emotional processing, consistent with the broader framework of stress-induced prefrontal and limbic dysregulation.

## 2. MATERIAL AND METHODS

### 2.1 Participants

This study was part of the VELAS (Ventral language stream in schizophrenia with regard to semantic and visuo-spatial processing anormalities)^42–45^. The sample (*N=*120, aged 17-39) comprised three groups matched for age and education: 40 patients with a first-episode SSD; within five years of onset), 40 individuals with high schizotypy (HS), and 40 with low schizotypy (LS).

SSD diagnoses were confirmed according to ICD-10 criteria: schizophrenia (F20; *N*=22), acute and transient psychotic disorder (F25; *N*=10), schizoaffective disorder (F25; *N*=5), and severe depression disorder with psychotic symptoms (F33.3; *N*=3). The 80 healthy controls were selected from a large-scale online screening of 1062 responders. All responders provided demographic information, answered questions regarding MRI safety, and completed two validated schizotypy scales: the Oxford-Liverpool inventory of feelings and experiences (O-LIFE)^46^ and the Multidimensional schizotypy scale (MSS)^47^. All responders and participants were required and to be native German speakers or possess native-level fluency, as all neuropsychological assessments were conducted in German. To ensure the healthy control group represented the same range of traits seen in the clinical sample, we employed a cluster-matching procedure. After identifying distinct clusters within the O-LIFE and MSS screening data using the *mclust* R package^48^, we applied the *predict.Mclust* function to the patient data. This allowed us to map the 40 SSD patients onto the established healthy clusters, revealing that the clinical profiles aligned consistently with the highest-scoring tiers. Consequently, the HS group was formed by selecting 40 healthy individuals from these same high-scoring clusters, while the LS group comprised 40 individuals from the lowest-scoring clusters.

Exclusion criteria for all participants were a current substance use disorder (excluding nicotine), a history of neurological or ophthalmological disorders, past head trauma involving loss of consciousness, and current pregnancy. Ethical approval was granted by the Cantonal Ethics Commission Zurich (KEK-ZH 2020/01049), and all participants provided written informed consent.

### 2.2 Neuropsychological assessment

All participants underwent a psychological assessment on the day of testing to provide a current psychopathological profile. This involved an evaluation of schizotypal personality traits via O-LIFE and MSS, alongside the administration of the Positive and Negative Syndrome Scale (PANSS)^49^.

The Childhood Trauma Questionnaire (CTQ)^50^, was administered to quantify various dimensions of early-life maltreatment, including emotional, physical, and sexual abuse, as well as emotional and physical neglect. To investigate potential cognitive impairments, participants completed a diverse battery of neuropsychological tests focusing on executive functions, memory and attention. Inhibitory control and automated response suppression was measured using the Victoria Stroop Test^51^. The Trail Making Test^52^ (parts A and B (TMT-A and B)) was employed to evaluate processing speed, cognitive flexibility, task switching and visual attention. Short-term and working memory were examined with the Digit-Span Forward and Backward respectively^53^.

Other high-order cognitive processes such as abstract reasoning, semantic flexibility and probabilistic reasoning were also measured through the administration of the Metaphor Triads Task (MTT)^54^ and the Mental Dice Task (MDT)^55^. Finally, a global measure of verbal IQ and crystallised intelligence was obtained using the Mehrfachwahl-Wortschatz-Intelligenztest (MWT-B)^56^.

### 2.3 MRI data acquisition

Neuroimaging data were acquired on a Philips Achieva 3.0 T scanner equipped with a 32-channel SENSE head coil. For each participant, we obtained CE-certified sequences including high-resolution structural T1-weighted images (1 × 1 × 1 mm³) and T2-weighted images for neuroradiological reading.

T1-weighted data were processed using surface-based morphometry in FreeSurfer, following a standard, fully automated pipeline widely used in morphometric studies and characterised by good test–retest reliability^57^.

### 2.4 Statistical analyses

Group differences in clinical scores were assessed using Kruskal-Wallis H tests with Benjamini-Hochberg adjusted Dunn’s post-hoc contrasts. Monotonic trends were evaluated via the Jonckheere-Terpstra test (Z statistic). Neuroanatomical differences across 71 defined regions of interest (ROIs) (52 cortical thickness measures and 19 sub-cortical/brainstem volumes) were analysed using Type III ANCOVAs^57,58^. Models were adjusted for age and gender with total intracranial volume (TIV) included for sub-cortical volumes only^59,60^. To mitigate the risk of Type I errors inherent in high-dimensional testing, the false discovery rate (FDR) was applied (*q<*.05). Significant group effects were decomposed via estimated marginal means (EMMs), with pairwise significance thresholds subjected to the Bonferroni correction to maintain a conservative inferential alpha.

To identify which neuroanatomical regions and traumatic experiences were most characteristic of each group across the psychosis spectrum, a multinomial logistic regression was employed. Using the low-schizotypy group as a baseline, this multivariate approach allowed us to determine the relative contribution of specific brain structures and ACEs in defining the clinical and high-schizotypy profiles. This step provided a more nuanced inferential insight than univariate tests alone, as it accounted for the simultaneous influence of multiple predictors. Multicollinearity was assessed via variance inflation factors (VIF) to ensure model stability.

The “interactome”, defined here as the comprehensive network of conditional dependencies between brain structures and schizotypal traits, was mapped using a mixed graphical model (MGM) with L_1_ LASSO regularisation. The λ parameter was determined using the extended Bayesian information criterion (EBIC)^61^. Childhood trauma was integrated into the network model as a categorical moderator using the cut-off of the CTQ (i.e., score*<*35 means no traumatic experience (“No ACE”; score≥35 means presence of a traumatic experience (“ACE”). Node strength, betweenness centrality, expected influence (EI), and node-wise predictability (R^2^)^62^ were calculated. Network architecture was compared using the network comparison test (NCT) with 1000 Monte Carlo permutations to global and edge-specific invariance^63^.

Analyses and network plots were performed using R version 4.5.2. Bar charts were prepared using GraphPad Prims Version 8.0.1.

## 3. RESULTS

### 3.1 Sample characterisation and childhood trauma distribution

A summary of the demographic, neuropsychological and clinical data is provided in Table 1.

**Table 1:** summary of the demographic profile, neuropsychological performance, and clinical assessment scores for the low schizotypy, high schizotypy, and patient groups. Data are expressed as mean (standard deviation) or absolute number (percentage) where appropriate. The cut-off scores represent the thresholds beyond which individuals within the study’s age range are defined as falling outside the normative range of the reference population. Note: family history refers to the “presence of psychotic illness in first degree relatives”; sd = standard deviation; CHF = Swiss Francs; TMT = Trail Making Test; PANSS = Positive and Negative Syndrome Scale; MSS = Multidimensional Schizotypy Scale; O-LIFE = Oxford-Liverpool Inventory of Feelings and Experiences; MWTB = Mehrfachwahl-Wortschatz-Intelligenztest; CTQ = childhood trauma questionnaire.

| DEMOGRAPHICS | Cutoff score | Low schizotypy | High schizotypy | SSD group |
| --- | --- | --- | --- | --- |
| **Sample size** | \ | 40 | 40 | 40 |
| **Gender** |  |  |  |  |
| - Males | \ | 18 (45%) | 21 (52.5%) | 28 (70%) |
| - Females | \ | 22 (55%) | 19 (47.5%) | 12 (30%) |
| Age | \ | 25.1 (sd 3.7) | 27.4 (sd 4.6) | 27.2 (sd 5.7) |
| Age of onset | \ | \ | \ | 24.7 (sd 6.3) |
| Years of education | \ | 14.5 (sd 2.3) | 14.8 (sd 2.4) | 13.3 (sd 3.1) |
| **Partnership** |  |  |  |  |
| - With a partner | \ | 24 (60%) | 25 (62.5%) | 7 (17.5%) |
| - Without a partner | \ | 16 (40%) | 15 (37.5%) | 33 (82.5%) |
| **Family history of mental disorder** |  |  |  |  |
| - Negative | \ | 38 (95%) | 34 (85%) | 33 (82.5) |
| - Positive | \ | 2 (5%) | 6 (15%) | 7 (17.5%) |
| **Neuropsychological assessment** |  |  |  |  |
| **Digit span** |  |  |  |  |
| - Forward (n) | ≤6(±1) | 7 | 6 | 6 |
| - Backward (n) | ≤(4±1) | 5 | 5 | 4 |
| **Trail making test** |  |  |  |  |
| - TMT-A seconds | ≥39 | 20.7 (sd 6.71) | 24.4 (sd 7.37) | 30.1 (sd 13.5) |
| - TMT-B seconds | ≥91 | 48.9 (sd 14.5) | 53.4 (sd 19.1) | 72.6 (sd 37.5) |
| **Victoria Stroop test** |  |  |  |  |
| - Error interference effect | \ | 0.21 (sd 0.40) | 0.12 (sd 0.46) | 0.51 (sd 0.73) |
| - Time interference effect | ≥12 | 5.61 (sd 2.76) | 5.82 (sd 2.95) | 9.65 (sd 6.12) |
| **Mental dice task (errors)** | \ | 0.1 (sd 0.3) | 0.22 (sd 0.4) | 1.08 (sd 1.47) |
| **MWTB-IQ** | ≤85 | 96.6 (sd 8.4) | 97.3 (sd 9.06) | 93.3 (sd 6.52) |
| **CTQ** | ≥35 | 30.06 (sd 5.39) | 35.32 (sd 9.67) | 42.3 (sd 15.44) |
| **Clinical assessment** |  |  |  |  |
| **PANSS** |  |  |  |  |
| - Positive | \ | 7.40 (sd 1.6) | 8.15 (sd 1.49) | 14.20 (sd 5.61) |
| - Negative | \ | 8.10 (sd 1.75) | 7.57 (sd 0.81) | 17.08 (sd 6.07) |
| - General psychopathology | \ | 16.6 (sd 2.77) | 17.0 (sd 2.42) | 30.50 (sd 6.75) |
| - Total score | ≥58 | 32.12 (sd 5.35) | 32.75 (sd 3.66) | 61.72 (sd 14.7) |
| **MSS** |  |  |  |  |
| - Positive | \ | 0.05 (sd 0.22) | 1.75 (sd 1.97) | 7.70 (sd 6.30) |
| - Negative | \ | 1.20 (sd 1.38) | 3.63 (sd 2.73) | 6.43 (sd 5.45) |
| - Disorganised | \ | 0.15 (sd 0.42) | 2.38 (sd 4.02) | 7.35 (sd 6.83) |
| - Total score | \ | 1.4 (sd 1.4) | 7.75 (sd 6.07) | 21.47 (sd 14.66) |

Table 1: summary of the demographic profile, neuropsychological performance, and clinical assessment scores for the low schizotypy, high schizotypy, and patient groups.
| O-LIFE |  |  |  |  |
| --- | --- | --- | --- | --- |
| - Cognitive disorganisation | \ | 0.75 (sd 0.87) | 7.68 (sd 5.57) | 11.5 (sd 6.23) |
| - Introverted anhedonia | \ | 1.30 (sd 0.88) | 4.13 (sd 2.67) | 7.58 (sd 5.65) |
| - Unusual experiences | \ | 1.25 (sd 1.37) | 5.2 (sd 4.22) | 10.1 (sd 6.98) |
| - Impulsive nonconformity | \ | 5.15 (sd 2.75) | 7.55 (sd 3.34) | 8.18 (sd 3.86) |
| - Total score | \ | 8.45 (sd 4.08) | 24.57 (sd 11.01) | 37.3 (sd 16.45) |

Neuropsychological comparisons revealed no significant differences in crystallised intelligence or memory capacity between groups. However, patients with SSD exhibited significantly slower processing on the TMT-A (*p<*0.001 vs LS) and TMT-B (*p<*0.001 vs LS; *p=*0.019 vs HS). In the Victoria Stroop Test the SSD group showed significantly higher time interference than the HS (*p=*0.006) and LS (*p=*0.002) groups. As expected clinical evaluation (PANSS, MSS, and O-LIFE) confirmed a significant stepwise increase in symptom severity across the psychosis spectrum(SSD > HS > LS), validating the group stratification (*p<*0.0001).

The most interesting trend was observed in the prevalence and burden of childhood adversity. Childhood trauma prevalence significantly increased alongside clinical severity: 32.5% in the LS group, 65% in the HS group, and 87.5% in the SSD group (*χ^2^*=25.88, *p<*0.001) (Figure 1). This linear gradient was further evidenced by total CTQ scores (*H=*30.16, *Z=*5.61; *p<*0.001) and the number of ACE categories reported (*H=*35.66, *Z=*5.87; *p<*0.001). Group disparities were most pronounced for physical neglect (*χ^2^*=23.27, *p<*0.001), emotional abuse (*χ^2^*=21.35, *p*=0.002), and emotional neglect (*χ^2^*=20.29, *p=*0.002). The prevalence of sexual abuse, likewise increased significantly across the cohort (*χ^2^*=17.09, *p=*0.009). Despite showing the lowest overall incidence, physical abuse was significantly differentiated between groups (*χ^2^*=9.7, *p<*0.046).

**Figure 1:**
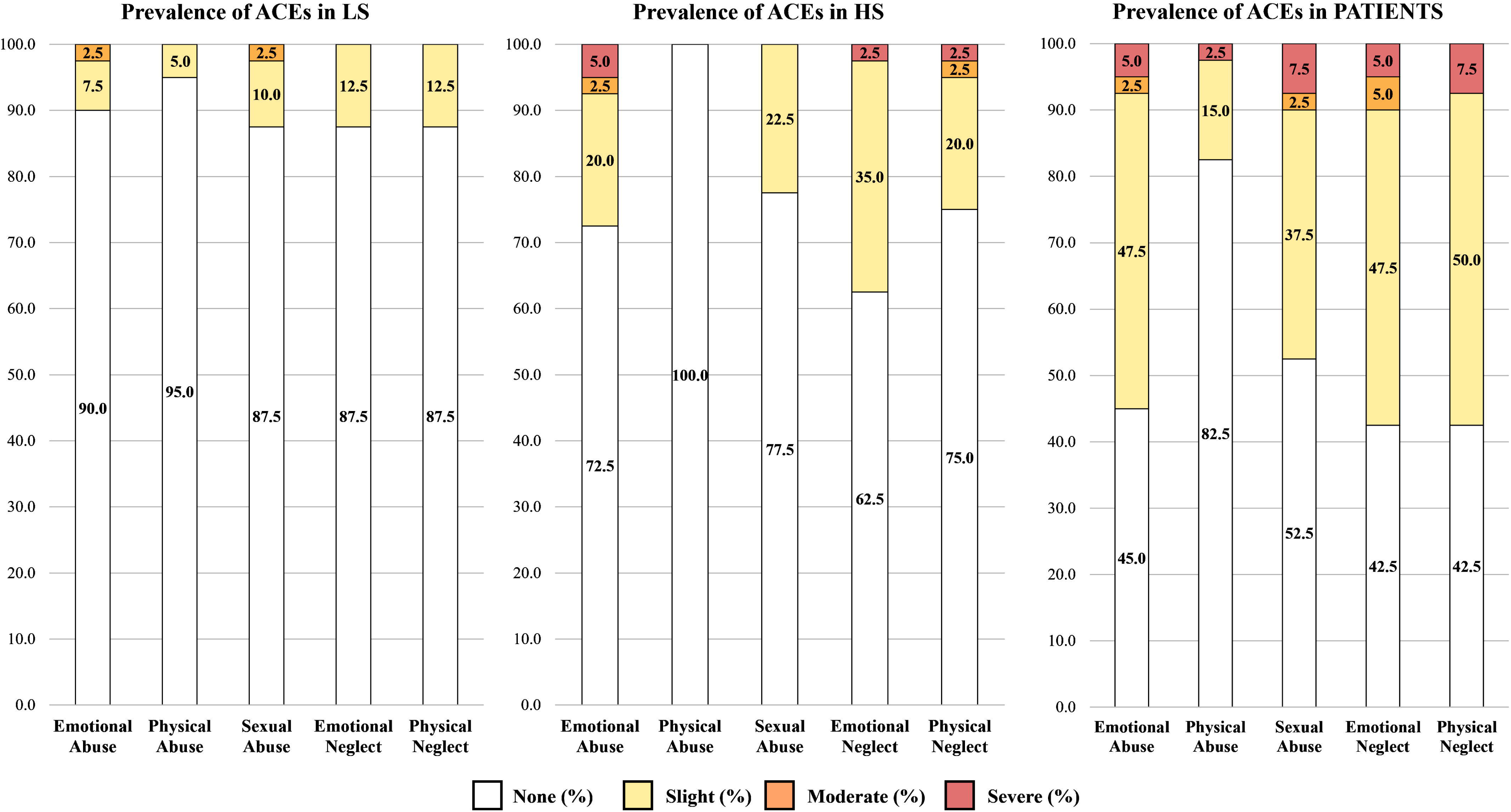
Distribution of Adverse Childhood Experiences (ACEs). Stacked bar charts illustrating the prevalence and intensity of ACEs across the three study groups: low schizotypy (LS), high schizotypy (HS), and patients with schizophrenia spectrum disorder (SSD). Levels of intensity (None, Slight, Moderate, and Severe) are derived from standardised scoring of the Childhood Trauma Questionnaire. Numerical values within each segment indicate the percentage of participants categorised at each intensity level within their respective group.

### 3.2 Neuroanatomical morphometry and regression results

Significant differences for the regions of interest (ROIs) are detailed in Figure 2. Notably, these structural differences are predominantly localised within a cohesive limbic-temporal-frontal network (Figure 2A). Pairwise contrasts revealed that 12 of 15 significant neuroanatomical differences were between the SSD and HS groups (Figure 2C). At the subcortical level, significant volumetric differences were exclusively observed between the HS and SSD groups, specifically localised within the bilateral amygdala and the left nucleus accumbens (Figure 2B). Within this network, the right posterior cingulate cortex (rPPC) emerged as the most robust anatomical marker, remaining the only region to maintain statistical significance across all group comparisons after FDR correction (*p_FDR_=*0.029).

**Figure 2:**
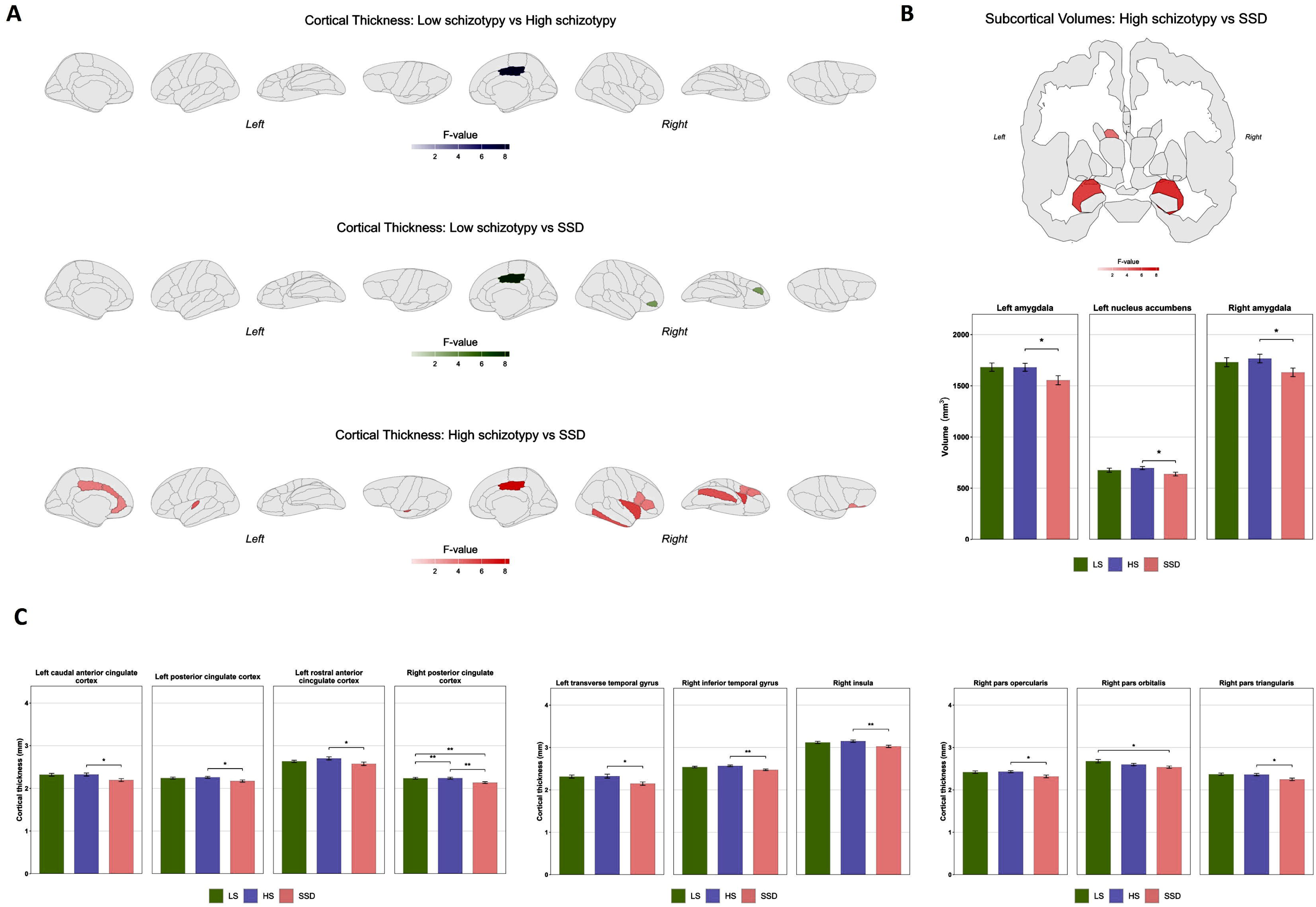
Group differences in cortical thickness and subcortical volumes. (A) Statistical parametric maps displaying significant regional differences in cortical thickness across three contrasts: Low Schizotypy (LS) vs High Schizotypy (HS), Low Schizotypy vs Schizophrenia Spectrum Disorder (SSD), and High Schizotypy vs SSD. Coloured regions indicate areas with significant differences (Bonferroni *p<*0.05), with the colour intensity representing the F-value. (B) Coronal bain slice to represent subcortical volume differences between HS and SSD patients (top). The associated bar charts (bottom) illustrate mean volumes (mm^3^) for these regions across the three study groups (LS, HS, and SSD). (C) Detailed regional bar charts for cortical thickness (mm). Significant differences (Bonferroni corrected) between groups are indicated by asterisks (*p<0.05, **p<0.01). Error bars represent the standard error of the mean (SEM).

To determine which of these structural markers and ACE scores independently characterise the psychosis spectrum, we employed a multinomial logistic regression (Baseline: Low Schizotypy; Figure 3). All variables returned VIF values<2.8 (Mean VIF=1.9), ensuring the stability of the estimated coefficients and the independence of the predictive contributions from bilateral regions.

**Figure 3:**
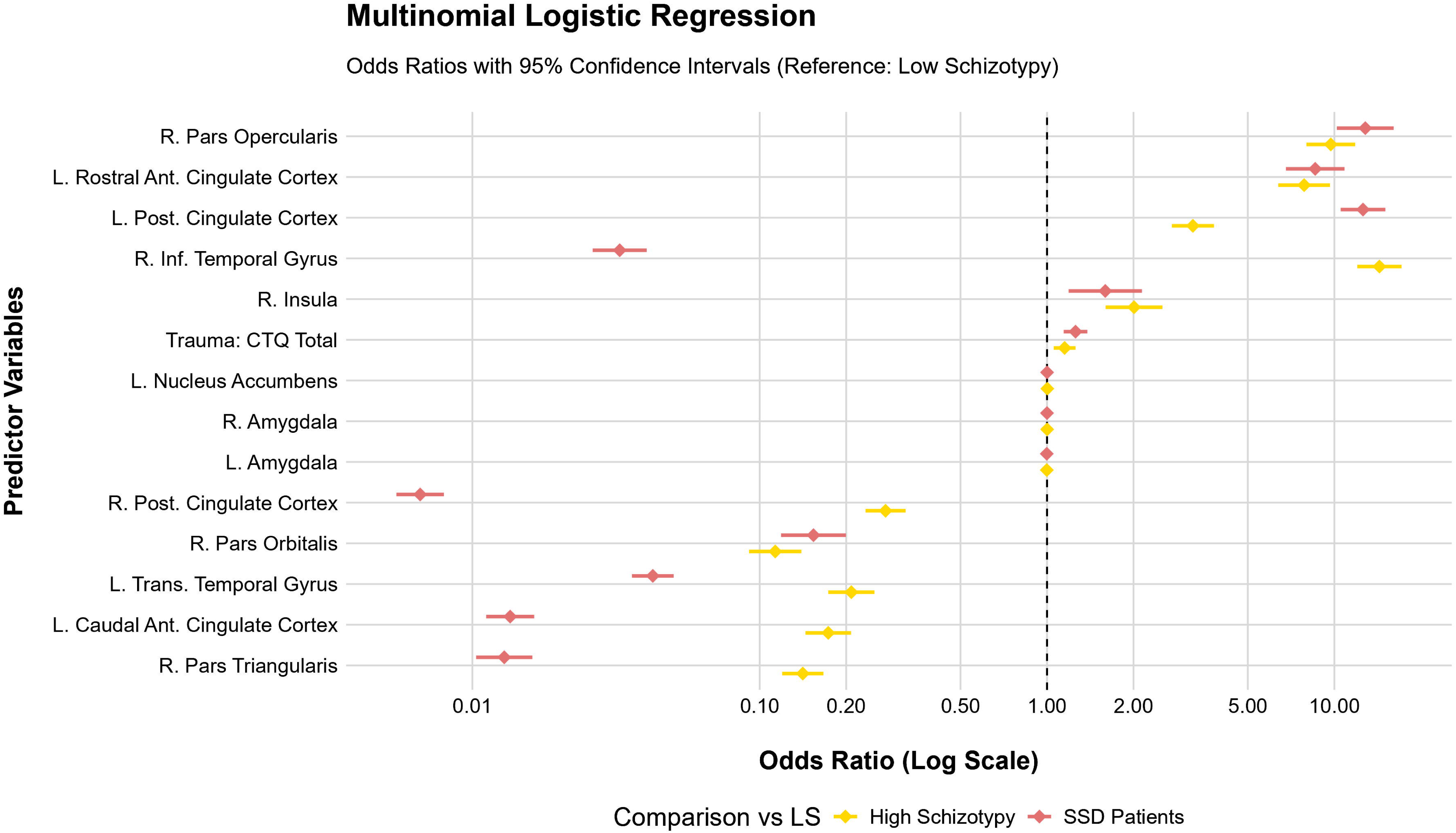
Forest plot displaying the odds ratios and 95% confidence intervals for variables predicting membership in the high schizotypy (yellow) and schizophrenia spectrum disorder (SSD; red) groups, using Low Schizotypy (LS) as the reference category (dashed vertical line, null effect). Values to the right (odds ratio > 1): Indicate that an increase in the predictor variable increases the likelihood of belonging to the HS or SSD group. Values to the left (odds ratio < 1): Indicate that a decrease in the predictor variable increases the likelihood of belonging to that group (i.e., higher values are associated with the reference LS group).

ACEs were associated with for both clinical and sub-clinical states: for every unit increase in the CTQ total score, the likelihood of belonging to the HS group increased by 15% (*OR=*1.15, *p=*0.0016), while the odds of belonging to the SSD group rose by 26% (*OR=*0.16, *p<*0.001). The multivariate model revealed two distinct structural patterns associated with increasing clinical severity:

Reduced cortical thickness in the right posterior cingulate cortex (rPCC; SSD: *OR=*0.006; HS: *OR=*0.27), left transverse temporal gyrus (lTTG; SSD: *OR=*0.04; HS: *OR=*0.20), and left caudal anterior cingulate (cACC; SSD: *OR=*0.01; HS: *OR=*0.17) were independent markers of membership in both the HS and SSD groups. Notably, the effect was significantly more pronounced in the SSD group, suggesting a progressive structural decline in these regions. Conversely, increased thickness in the left rostral anterior cingulate (left rACC; SSD: OR=8.58; HS: OR=7.85), the right pars opercularis (SSD: OR=12.80; HS: OR=9.72), and the right insula (SSD: OR=1.60; HS: OR=2.01) characterised the pathological groups.

Notably a unique profile was observed for the right inferior temporal gyrus (rITG). While increased thickness in this region was strongly linked to the HS phenotype (*OR=*14.35, *p<*0.001), reduced thickness was associated with the SSD group (*OR=*0.03, *p<*0.001), potentially marking a neurobiological divergence point between sub-clinical and clinical states. Within this multivariate framework, subcortical volumes (amygdala and nucleus accumbens) did not provide additional explanatory value beyond the cortical markers (*p>*0.05).

### 3.3 Moderated network dynamics

Overall, childhood trauma appeared to fundamentally reorganise the brain-personality network. At a structural level, early adversity increased network density and elevated the rostral anterior cingulate cortex as a central hub, with strengthened links to the insula and amygdala. At the brain-behaviour level, in the trauma group the network’s focus moved away from typical cognitive-executive schizotypy and towards affective and impulsive dimensions, anchored by strengthened links to the amygdala. These results suggest that trauma shifts the brain-behaviour system from a cognitive orientation toward a limbic-dominant organisation.

#### 3.3.1 Structural network reconfiguration

Specifically, the moderated mixed graphical model, constructed using the nominally significant brain regions and ACEs as a moderator, revealed a reconfiguration of the structural covariance interactome. As illustrated in Figure 4, the network topology for the ACE group (part B) exhibited a significant higher connectivity density (37.36%) compared to the No ACE group (Part A, 15.38%). Formal testing via the NCT confirmed a significant difference in global network architecture between the two conditions (network invariance *M=*0.383, *p=*0.03), while no significant difference was observed in global strength (*S=*0.137, *p=*0.88).

**Figure 4:**
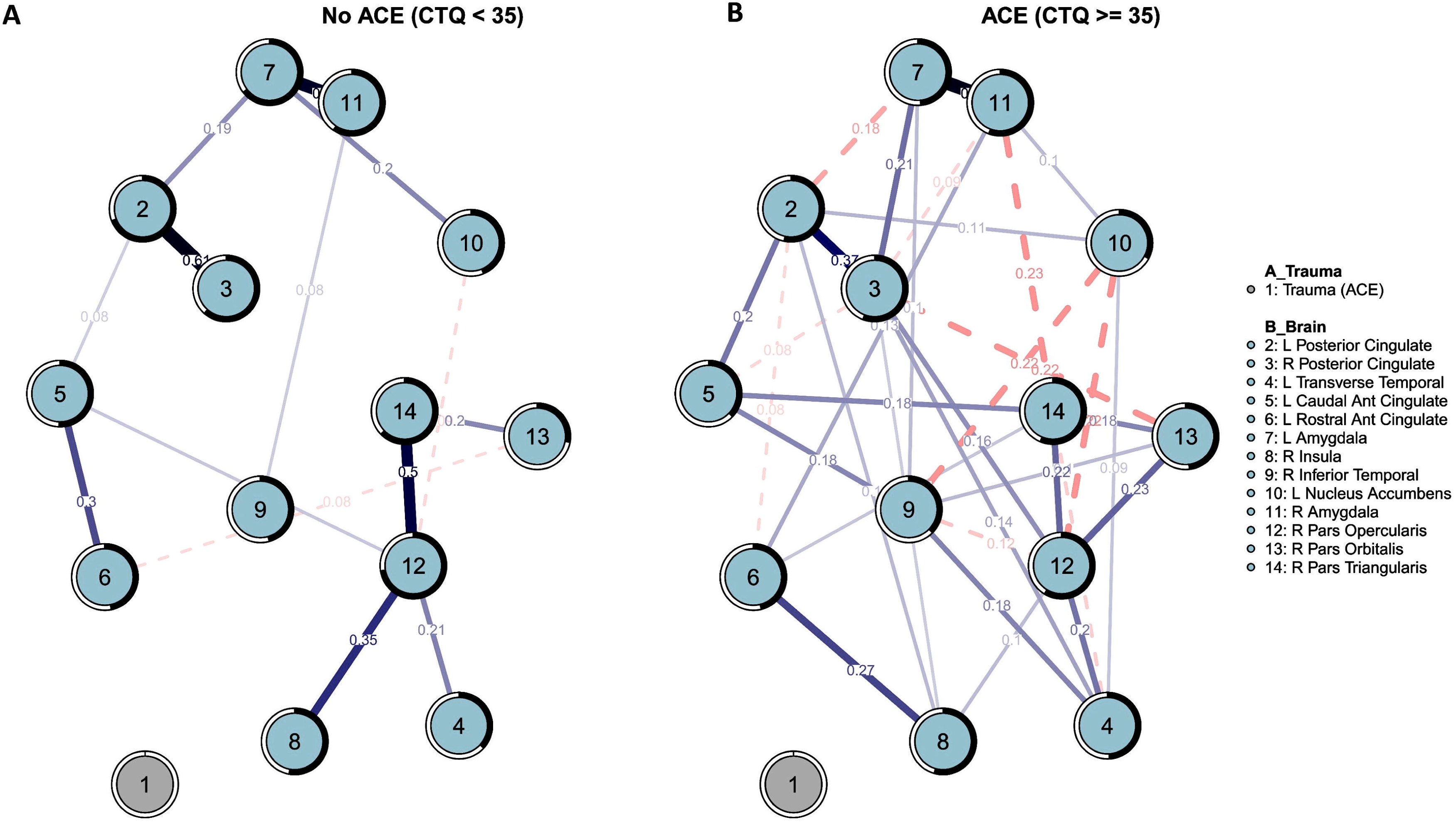
Comparison of conditioned brain networks representing individuals with (A) no adverse childhood experiences (low trauma: CTQ < 35) and (B) positive history of adverse childhood experiences (high trauma: CTQ ≥ 35). Edges represent the partial correlation between regions after conditioning on the moderator. Blue solid lines indicate positive associations, while red dashed lines represent negative associations. The thickness and saturation of the edges are proportional to the strength of the connection (edge weights). Note: ACE (adverse childhood experience), CTQ (childhood trauma questionnaire).

Specific edge invariance tests identified the key pathways targeted by this rewiring. The most robust difference occurred in the link between the left rACC and right insula (*p=*0.01). Significant shifts were also noted in the covariance between the anterior cingulate (left cACC and the left rACC; *p=*0.03), the inter-hemispheric connection of the default mode network (left to right posterior cingulate; *p=*0.05), and the dependence between left cACC and lITG (*p=*0.03). Furthermore, trauma altered the associations between the left rACC and both the right amygdala (*p=*0.04) and the right pars triangularis (*p=*0.04).

Centrality invariance tests further highlighted a shift in the topological role of the left rACC, which saw its betweenness centrality nearly double (0.08 to 0.15, *p=*0.04). While the right pars opercularis a primary hub across both groups, the rPCC emerged as the primary hub of strength centrality in the ACE condition (relative strength: 1.00 compared to 0.49 in the No ACE group).Node-wise predictability remained robust, though the ACE group displayed slightly lower predictability for limbic nodes like the left amygdala (R^2^ decreased from 0.646 to 0.486) and the left nucleus accumbens (R^2^ decreased from 0.439 to 0.328).

#### 3.3.2 Brain-schizotypal personality interactome

Incorporating schizotypy dimensions (MSS and O-LIFE) revealed a comprehensive interactome where trauma moderated the bridge between biology and personality (Figure 5). While global strength was stable (*S=*0.27, *p=*0.77), the NCT indicated a trend toward global architecture reconfiguration (*M=*0.4, *p=0.07*)

**Figure 5:**
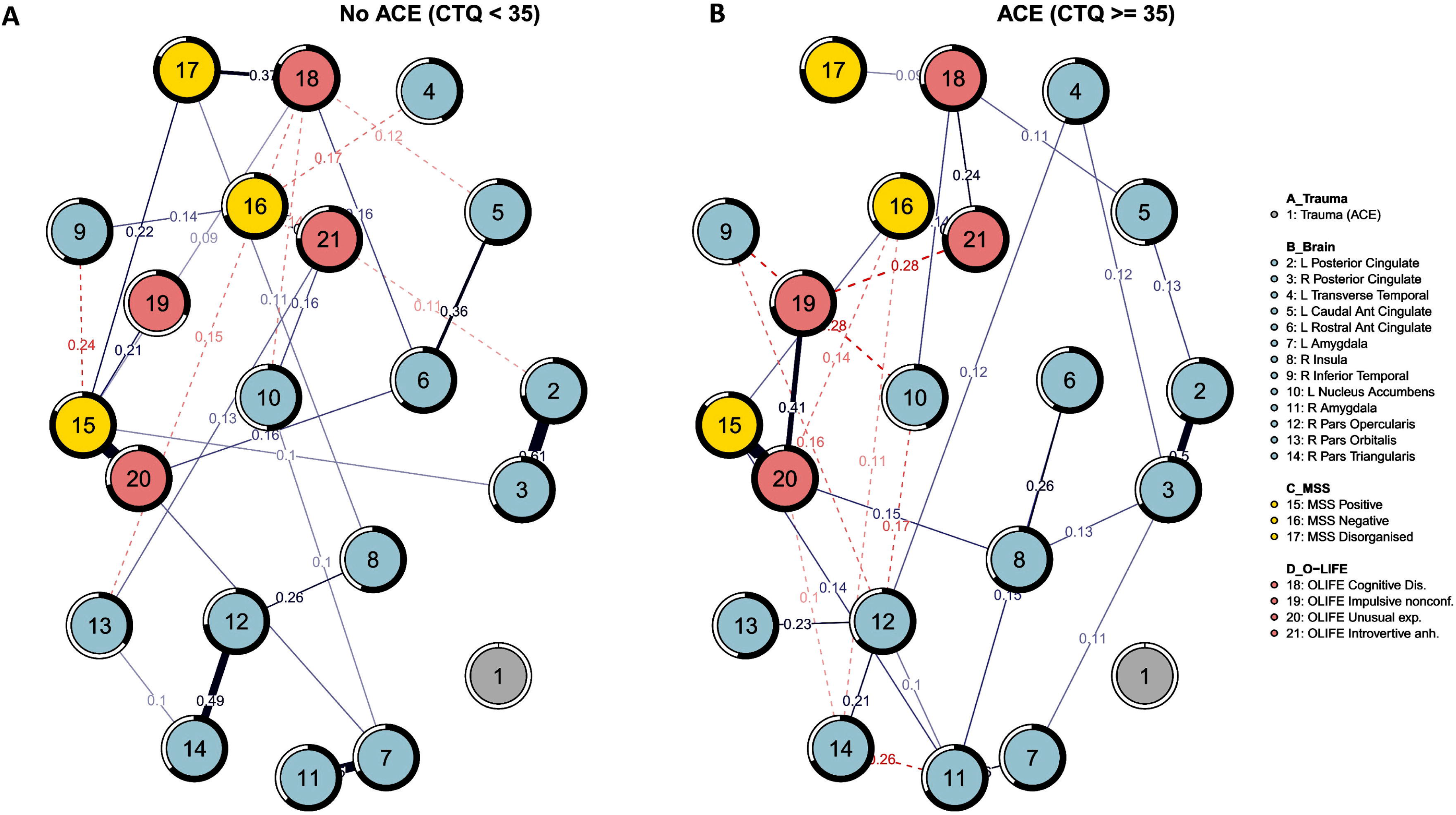
Comparison of conditioned brain networks representing individuals with (A) no adverse childhood experiences (low trauma: CTQ < 35) and (B) positive history of adverse childhood experiences (high trauma: CTQ ≥ 35). Edges represent the partial correlation between regions after conditioning on the moderator. Blue solid lines indicate positive associations, while red dashed lines represent negative associations. The thickness and saturation of the edges are proportional to the strength of the connection (edge weights). Note: ACE (adverse childhood experience), CTQ (childhood trauma questionnaire).

Several pathways were significantly moderated by trauma: while brain-brain covariances remained robust (left and right PCC,*p=*0.031; left cACC and left rACC, *p=*0.014; left rACC and the right insula, *p=*0.007), trauma significantly altered the relationship between the MSS positive subscale and key regions including the rACC (*p=*0.013) and the right amygdala (*p=*0.046). A particularly marked impact of the trauma moderation emerged for the O-LIFE impulsive nonconformity scale. Beyond an increase in predictability (from R^2^=0.31 in the No ACE group to 0.73 in the ACE group), this schizotypy component revealed significantly altered edge invariance with the left amygdala (*p=*0.006). Similarly, the O-LIFE introvertive anhedonia dimension showed reconfigurations in its links with left amygdala (*p=*0.006), and right amygdala (*p=*0.037).

Centrality invariance tests showed that the EI of the MSS positive subscale differed significantly (*p=*0.002); while it was the most influential node in the No ACE group (1.00), its influence decreased in the trauma group (0.64), where O-LIFE unusual experiences emerged as the primary hub of network (relative EI = 1.00). Notably, the O-LIFE impulsive nonconformity subscale showed a tripling of its relative influence (increasing from 0.14 to 0.45. The O-LIFE unusual experiences subscale showed the highest predictability in the ACE group (*R^2^=*0.903), while the O-LIFE impulsive nonconformity node showed the most dramatic increase (*R^2^* difference=+0.419). Conversely, structural nodes generally showed slight declines in predictability in the ACE group, most notably the lPCC (No ACE *R^2^=*0.72; ACE *R^2^=*0.60) and the rITG (No ACE *R^2^=*0.58; ACE *R^2^=*0.46).

In summary, childhood trauma reconfigured the brain-personality network toward limbic dominance, with the anterior cingulate and amygdala emerging as pivotal nodes linking structural vulnerability to the affective and impulsive dimensions of schizotypy.

## 4. DISCUSSION

Childhood trauma has long been recognised as a primary environmental risk factor for the development of psychosis, yet the systemic mechanisms through which early adversity reconfigures the interactions between brain and personality (which we refert to as interactome in this study) remain poorly understood. This study aimed to address this gap by combining morphometric analyses with network theory to examine the interplay between adverse childhood experiences, brain structure, and multidimensional schizotypal traits along the psychosis spectrum. We found that childhood trauma reorganised the structure of psychosis risk, shifting the brain-personality network toward limbic dominance and amplifying affective dimensions of schizotypy. Clinically, this pattern suggests that trauma may reduce network resilience by strengthening vulnerability hubs and amplifying the influence of affective schizotypy dimensions, creating a configuration in which local perturbations are more likely to propagate into broader psychopathological expressions.

### 4.1 The traumatogenic phenotype and social defeat

In alignment with the first two hypotheses, we observed a clear, stepwise increase in ACE prevalence across the studied groups (LS<HS<SSD). This provides empirical support for the “traumatogenic phenotype” model, which posits that certain clinical profiles are shaped by the developmental impact of early trauma, and extends its validity to the subclinical domain^64^. Notably, the high prevalence of trauma in the clinically healthy HS group, double that in the LS group, supports the conceptualisation of schizotypy as a personality-level manifestation of this same traumatogenic vulnerability^65,66^. These findings also align with the “social defeat hypothesis” which is derived from the vulnerability-stress model. This hypothesis suggests that chronic exposure to adverse, or abusive environments acts as “sensitising agent” for the brain’s stress response system^16^. By experiencing early-life neglect or abuse, individuals may undergo a permanent shift in their “threat-detection” threshold. This results in neutral social stimuli being increasingly interpreted through a lens of suspicion or withdrawal, a hallmark of schizotypal traits^67^. Our complex systemic analysis provides insights into how this psychological state relates to a cohesive neuroanatomical fingerprint that reconfigures the structural covariance of the brain.

### 4.2 The rPCC as a vulnerability hub

Our results indicate that systemic sensitisation appears to be concentrated within a specific structural interactome comprising temporal, frontal, and limbic regions. The rPCC emerged as the most robust indicator of phenotypic severity, exhibiting a significant stepwise reduction in cortical thickness across the three groups (LS>HS>SSD). This finding is critical when viewed through our network analysis: the rPCC became the primary hub of strength centrality only in the presence of trauma.

As a core node of the default mode network (DMN), the rPCC is fundamental to self-referential processing and the integration of internal versus external information^68^. The differences observed in our sample suggest that trauma exposure is linked to a lower structural integrity of this hub, centralising the system’s weight onto a single, thinning region. We hypothesise that such a structural shift might diminish the system’s capacity to flexibly process social information, potentially anchoring the individual in the self-referential ruminations and social withdrawal typical of the psychosis spectrum^69^.

### 4.3 Structural decoupling and executive-salience disruption

The vulnerability in the rPCC is matched by a significant imbalance in the ACC. While thinness in the left cACC predicted membership of the SSD group, greater thickness in the left rACC was indicative of both HS and SSD phenotypes. This data suggests a breakdown in the coordinate maturation of the ACC^70^. The NCT revealed that the covariance between these two regions is significantly reconfigured, potentially disrupting the top-down regulation of emotional salience.

Two other areas, the rITG and the lTTG, showed a more complex profile: whereas reduced thickness in the lTTG and rITG predicted SSD, increased thickness in the rITG was a powerful predictor of the HS phenotype^19,71^. This finding suggests that in non-clinical individuals, a state of compensatory hypertrophy may exist wherein healthy HS individuals recruit temporal resources to maintain cognitive coherence despite their underlying liability^72,73^. However, network results show that childhood trauma is associated with a reduction in the node-wise predictability of the rITG. This indicates that its statistical dependencies with other regions in the system become less informative, suggesting that trauma may shift the drivers of rITG morphology toward other external factors. Finally, the predictive model highlighted a clear atrophy-driven progression in the right pars orbitalis and pars triangularis (i.e., inferior frontal gyrus)^74^. These regions are critical for executive control and the inhibition of inappropriate responses.

In the context of the salience network reconfiguration, our findings point toward a structural breakdown of top-down regulatory pathways. This is consistent with neurodevelopmental models suggesting that early adversity triggers an affective-cognitive decoupling, where atrophied prefrontal regions fail to provide the necessary morphological substrate for the effective modulation of stress-sensitive salience circuits^75,76^.

### 4.4 Limbic anchoring of schizotypy personality traits

This physical weakening of the brain’s regulatory centres is reflected in a fundamental shift in what drives schizotypal personality. In individuals without a history of trauma, schizotypy is primarily defined by cognitive and perceptual traits (MSS positive subscale acts as the most influential node). However, exposure to trauma shifts this balance, changing which traits dominate the network and how they interact. In the ACE group the O-LIFE impulsive nonconformity subscale emerged as a central driver, with its relative EI increasing from 0.14 to 0.45. This shift suggests that early-life adversity anchors schizotypal traits within subcortical emotional centres, transforming schizotypy from a predominantly cognitive-perceptual vulnerability to one that also includes an affective-behavioural dimension.

The NCT results clarify this transition: in the ACE network, trauma alters the underlying connections between nodes, creating a different set of dependencies. The impulsive nonconformity and introvertive anhedonia dimensions, reveal significantly altered edge invariance with the amygdala and the nucleus accumbens. This is matched by an increase in the predictability of impulsive traits, indicating that in the presence of ACEs, these features become a central, highly integrated component of the brain’s structural-phenomenological architecture.

Collectively, these reconfigurations suggest that childhood trauma alters the network’s structure. While an interactome not subjected to ACEs is organised around core functioning personality dimensions, the trauma-exposed system is prioritised toward subcortical emotional reactivity. This “limbic anchoring” might explain why trauma-exposed individuals exhibit higher stress reactivity and more diverse clinical trajectories: their personality traits are no longer governed by stable cortical scaffolding but are instead hard-wired into the brain’s primary affective and reward centres^32,77^.

### 4.5 Limitations

Despite the robustness of our multivariate and network-based findings, several limitations must be acknowledged. First, our cross-sectional design precludes definitive conclusions regarding causality. Nevertheless, a model in which early trauma acts as the primary driver of cortical thinning and network shifts is consistent with neurodevelopmental theories and the traumagenic model of psychosis. Second, while our sample size (*N=*120) provided sufficient power for the primary group comparisons and regularised regressions, it may limit the generalisability of the more complex moderated network interactions, which would benefit from replication in larger, multi-site cohorts. Furthermore, the assessment of ACEs relied on the CTQ. Although it is the validated gold standard for assessing childhood trauma, it remains subject to retrospective recall bias. Finally, our model did not account for other environmental variables known to influence the psychosis spectrum, such as urbanicity, cannabis use, and current life stressors. These factors are known to interact with childhood adversity and could further moderate the brain–behaviour interactome, potentially explaining additional variance in our structural and phenomenological nodes.

### 4.6 Conclusion

In conclusion, this study offers a high-resolution map of the way ACEs act as a systemic catalyst within the dependencies between brain areas and schizotypal personality traits. The strength of this work lies in its integrative approach. Moving beyond a reductionist “one-region, one-symptom” paradigm, we were able to observe and explain complex relations between neuroanatomical, environmental, and phenomenological dimensions. Future research should seek to explore these interaction dynamics in greater detail from two perspectives: firstly, descriptively, by creating models for individual subgroups of individuals on the schizotypy spectrum, both for different types and intensities of childhood trauma; and secondly, temporally/causally, by employing longitudinal network models to obtain a clearer picture of how they evolve.

## Data Availability

All data produced in the present study are available upon reasonable request to the authors

## AKNOWLEDGEMENTS

This research was supported by grants from the Brain & Behaviour Research Foundation (Grant No. 28997, to W.S.), the OPO Foundation (Grant No. 2020-0075, to W.S. and P.H.), and the European Research Council (ERC Synergy; Grant No. 101118756, to P.H.). We gratefully acknowledge their financial assistance, which made this work possible.

